# “Prevalence, Patterns, Clinico-social and Behavioural factors associated with the Consumption of Sugar Sweetened Beverages among Undergraduate Medical Students of Central India”

**DOI:** 10.1101/2021.08.29.21262509

**Authors:** Soumya Kanti Mandal, G Revadi, Darshan Parida, Anindo Majumdar

## Abstract

**Background:** Excessive consumption of Sugar Sweetened Beverages (SSBs) in adolescents has become a global issue. As its link to obesity and non-communicable diseases is clear, it is imperative to understand SSB consumption behaviours in the future healthcare professionals.

**Objective:** To document the prevalence, patterns and clinic-social and behavioural factors predicting high intake of SSBs among medical students.

**Methods:** This cross-sectional study was conducted using a self-reported, web-based, questionnaire. All the students and interns who were part of a publicly funded premiere teaching hospital during October and November 2019 were included. The semi-structured questionnaire enquired regarding socio-demographic, clinical details, amount, behavioural patterns and money spent in connection with SSB consumption. Data were analysed using IBM SPSS version 24.

**Results:** The mean (SD) age of participants was 19.3 (1.6) years, 71.7% being males. The current prevalence of SSB consumption was 90.5 %. Also, 49.9% and 29.1% participants preferred soft drinks and sweetened fruit juice respectively. Multivariable analysis showed that male gender (aOR 1.83, 95% CI 1.03-3.25), current alcohol consumption (aOR 4.09, 95% CI 1.25-13.42), and recent (last week) consumption of a SSB predicted high intake of SSBs (aOR 7.36, 95% CI 3.41-15.87) whereas, preference of energy/sports category of drinks predicted low intake of SSBs (aOR 0.10, 95% CI 0.02-0.47).

**Conclusion:** The consumption of SSBs among medical students was high. Targeted health education and behaviour change interventions should be provided to males, alcohol users and frequent consumers.

## Introduction

Globally, in 2016, more than 1.9 billion adults were overweight. Of these, over 650 million were obese. (*Obesity and overweight*, no date a)Not only adults, adolescent obesity has also become a problem. In 2016, over 340 million in 5-19 age group were found to be overweight.(*Obesity and overweight*, no date b) Excessive consumption of Sugar Sweetened Beverages (SSBs) have been found to contribute to increase in prevalence of obesity, tooth decay and NCDs.(Malik, Schulze and Hu, 2006; Sohn, Burt and Sowers, 2006; Malik *et al*., 2010; Yang *et al*., 2014) SSBs are any liquids that are sweetened with various forms of added sugars like brown sugar, corn sweetener, corn syrup, dextrose, fructose, glucose, high-fructose corn syrup, honey, lactose, malt syrup, maltose, molasses, raw sugar, and sucrose.(CDC, 2018) Examples of SSBs include but are not limited to regular soda (not sugar-free), fruit drinks, sports drinks, energy drinks, sweetened waters, and coffee and tea beverages with added sugar.(CDC, 2018) SSBs have little nutrition value.(*WHO* | *Reducing consumption of sugar-sweetened beverages to reduce the risk of childhood overweight and obesity*, no date)

Since 1998, SSB sales in India have been increasing by 13% year-on-year exceeding 11 litres per capita per year.(Basu *et al*., 2014) According to a study based on National Family Health Survey, round 4 (NFHS 4) data, in the Indian population, the highest amount of consumption of SSBs has been seen among adolescents.(Mathur *et al*., 2020) In other parts of the world also, adolescents and young adults were the highest consumer of SSBs.(Singh *et al*., 2015) Among adolescents, being male, fast food consumption and watching TV were found to be associated with higher consumption of SSBs, but high physical activity are associated with low consumption.(Park *et al*., 2012a) In another study, rural living environment, less knowledge of energy expenditure were found to be associated with higher consumption.(Wattelez *et al*., 2019) Most of the undergraduate medical students are adolescents or young adults. If their health is not optimum, this will affect patient care. Also, medical professionals consuming SSBs themselves would not be seen as ideal role models by their patients, who usually look up to their treating physician.

Because of the above-mentioned reasons, patterns of SSB consumption and associated clinic-social and behavioural factors are important to understand. Previous studies from India have analysed secondary data, and also have studied few variables.(Mathur *et al*., 2020) Also, we could not find any study conducted among medical students.

The objectives of the present study were, thus, to document the prevalence and patterns of SSB consumption and to find out the clinic-social and behavioural factors predicting higher intake of SSBs among undergraduate medical students of a premiere tertiary care teaching hospital in central India.

## Methodology

This cross-sectional study was conducted among all the undergraduate medical students and interns of a premier tertiary care teaching hospital of central India who were part of this institute during October and November 2019. Students who were on long term leave due to any disease or were absent continuously for at least one month during the study period were planned to be excluded.

A web-based self-administered questionnaire was developed using the Kobo Toolbox (Harvard Humanitarian Initiative), which is free for non-commercial use.(*KoBoToolbox* | *Data Collection Tools for Challenging Environments*, no date) The semi-structured questionnaire was developed in English language having both closed and open-ended questions. It had questions regarding socio-demographic, clinical details, amount and patterns of SSB consumption, and the money spent for SSB consumption. We also asked the participants about the term ‘Sugar Sweetened Beverage’, to see if he/she had ever come across this term in media/scientific articles/textbooks, which might have also influenced their consumption patterns.

The class representatives were briefed about the study and a link to the questionnaire, along with the electronic copies of participant information sheet and a copy of consent form were shared with them on WhatsApp (Facebook Corp), which is a mobile messaging application. The students were then asked to fill their responses in the next 20 minutes. At least two reminders were sent to students (absentees and non-responders) through phone call, WhatsApp or personal contact before marking him/her as non-responder. Clearance from the institutional human ethics committee of AIIMS Bhopal was obtained (IHEC-LOP/2018/STS0146) prior to start of study.

The following operational definitions were used in the present study:

Current tobacco use was defined as tobacco used in any form, either smoked or smokeless, in the last one month. Current alcohol use was defined as any amount of alcohol consumed in the last one month. Sufficient physical activity (Yes/No) was defined as perceived self-reported amount of physical activity. Enough sleep (Yes/No) was defined as perceived self-reported amount of sleep.

SSB was defined as liquids sweetened with various forms of added sugars like brown sugar, corn sweetener, corn syrup, dextrose, fructose, glucose, high-fructose corn syrup, honey, lactose, malt syrup, maltose, molasses, raw sugar, and sucrose. SSBs include but are not limited to regular soda (not sugar-free), fruit drinks, sports drinks, energy drinks, sweetened waters, and coffee and tea beverages with added sugar.(CDC, 2018) For the purpose of our study, we limited this definition to beverages which were sold in the market and purchased by people, and not homemade beverages, like coffee, tea, etc. Current prevalence of SSB consumption was defined as the number of participants who had consumed any form of SSB in the last one month out of total number of participants included in the study. Lifetime prevalence of SSB consumption was defined as the number of participants who had ever consumed any form of SSB in their lifetime out of total number of participants included in the study.

High intake of SSB was defined as consumption of ≥ 1 litre (L) of any form of SSB/per participant/month. Last/recent consumption of SSB was defined as consumption of SSB in any form in the last week (seven days).

Education and occupation of parents were classified according to Modified Kuppuswamy scale.(Wani, 2019) For education, the original categories used for data collection were: profession or honours, graduate or post graduate, intermediate or post high school diploma, high school certificate, middle school certificate, primary school certificate and illiterate. For occupation, the categories used were: profession, semi-profession, clerical/shop owner/farmer, skilled, semi-skilled, unskilled and unemployed. During analysis, some of these categories were clubbed together to re-categorise as the numbers were small in original categories to draw any meaningful conclusions. Per capita income was calculated by dividing total family income by number of family members. For the sake of analysis in this study, it was further categorized as 0-68.72 USD, 68.74-343.67 USD, and >343.68 USD. This was further recategorized into <68.74 and ≥68.74 USD for further analysis.

Data were exported from Kobo Toolbox to Microsoft Excel 2010 and analysis was performed using Statistical Package for Social Sciences version 24 (IBM SPSS). Proportions and means were calculated, along with 95% confidence interval (CI) and standard deviation (SD), respectively. Chi-square test was used to compare proportions among groups and to test associations. Univariable logistic regression analysis was conducted to understand the predictors of increased SSB consumption, especially the socio-demographic and the clinical related factors among the participants. Unadjusted odds ratio was calculated. The variables which had a P value <0.25 in univariable analysis were entered in the multivariable model. Multivariable logistic regression analysis was performed to find out the independent predictors of increased SSB consumption and adjusted odds ratios were reported. *P* value <0·05 was considered to be significant.

## Results

A total of 358 responses were recorded using Kobo Toolbox out of 499 students who were sent the link to the questionnaire (28.3% non-response). Out of 358, one participant had missing data/inappropriate responses in most fields, so the total valid responses analysed were of 357 participants. None of the participants fit the exclusion criteria. Table 1 provides the details of sociodemographic and clinical characteristics of the participants. Out of 357, the majority (71.7%) were male, were below 20 years of age (59.9%), had permanent residence in an urban area (72.8%) and belonged to a nuclear family (85.7%). The median (IQR) per capita income per month was 183.29 (91.65-343.68) USD ranging from nil to 14663.8 USD. A total of three (0.8%) participants had history of chronic disease namely diabetes, hypertension and cardiovascular disease.

**Table 1:**
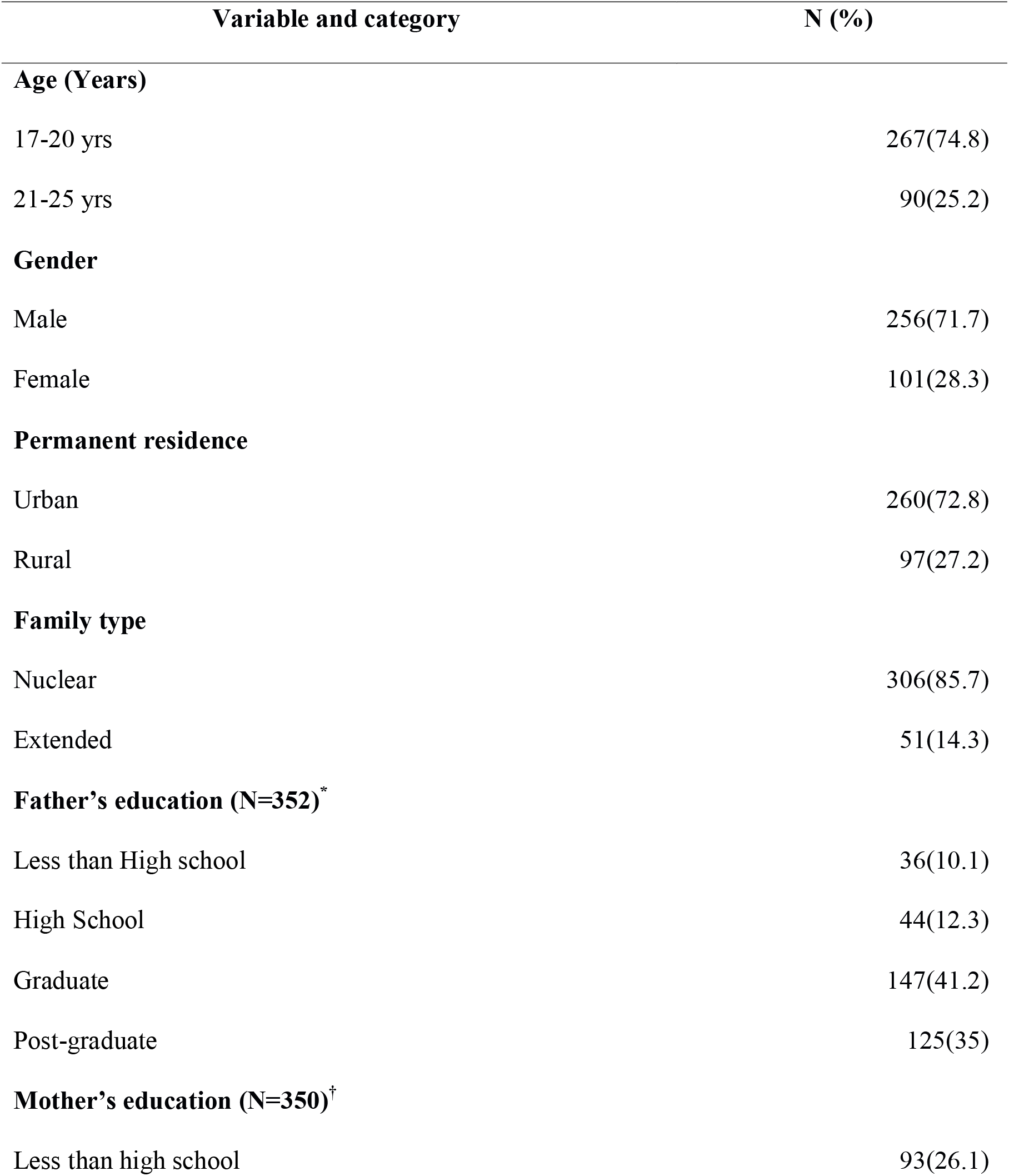
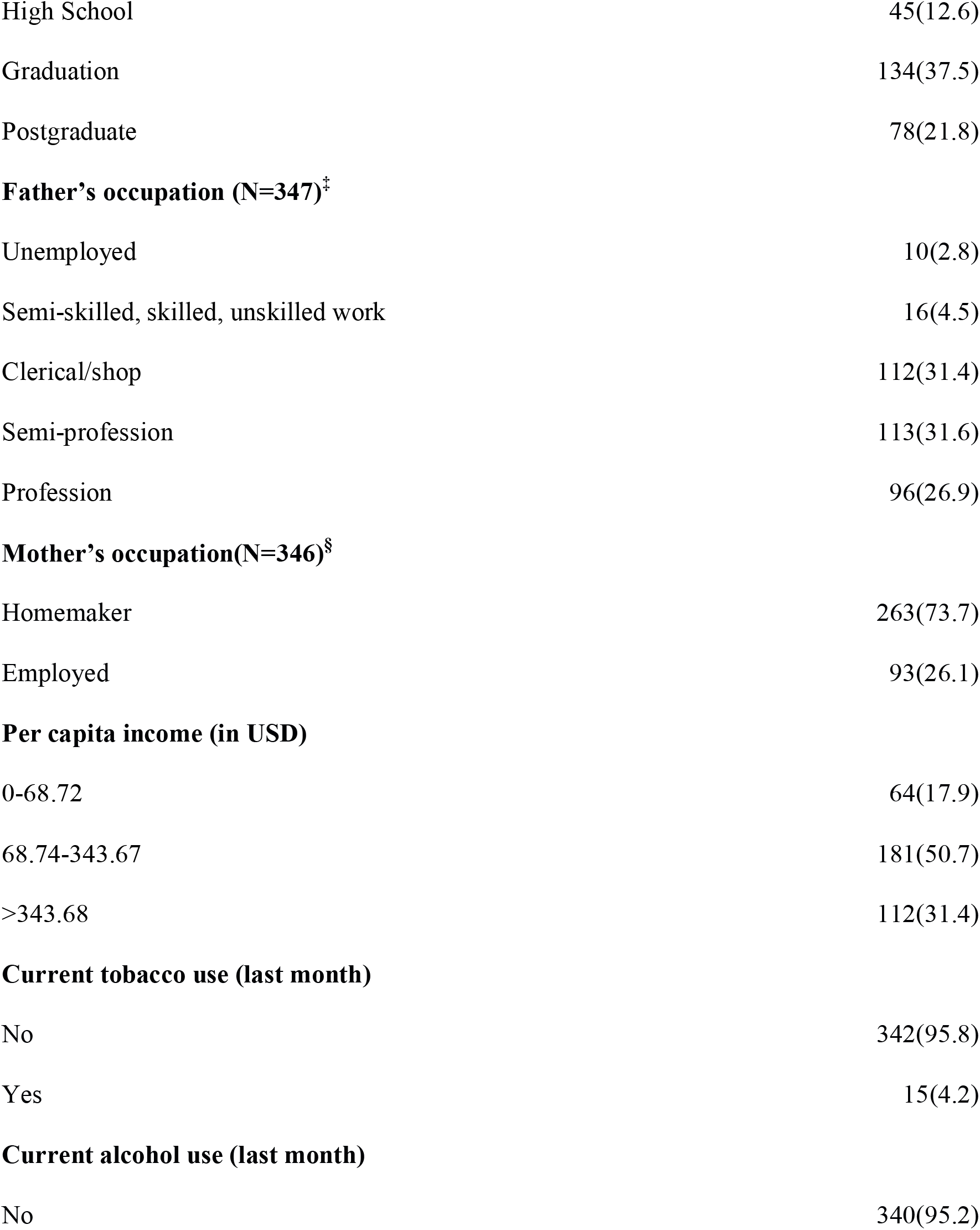
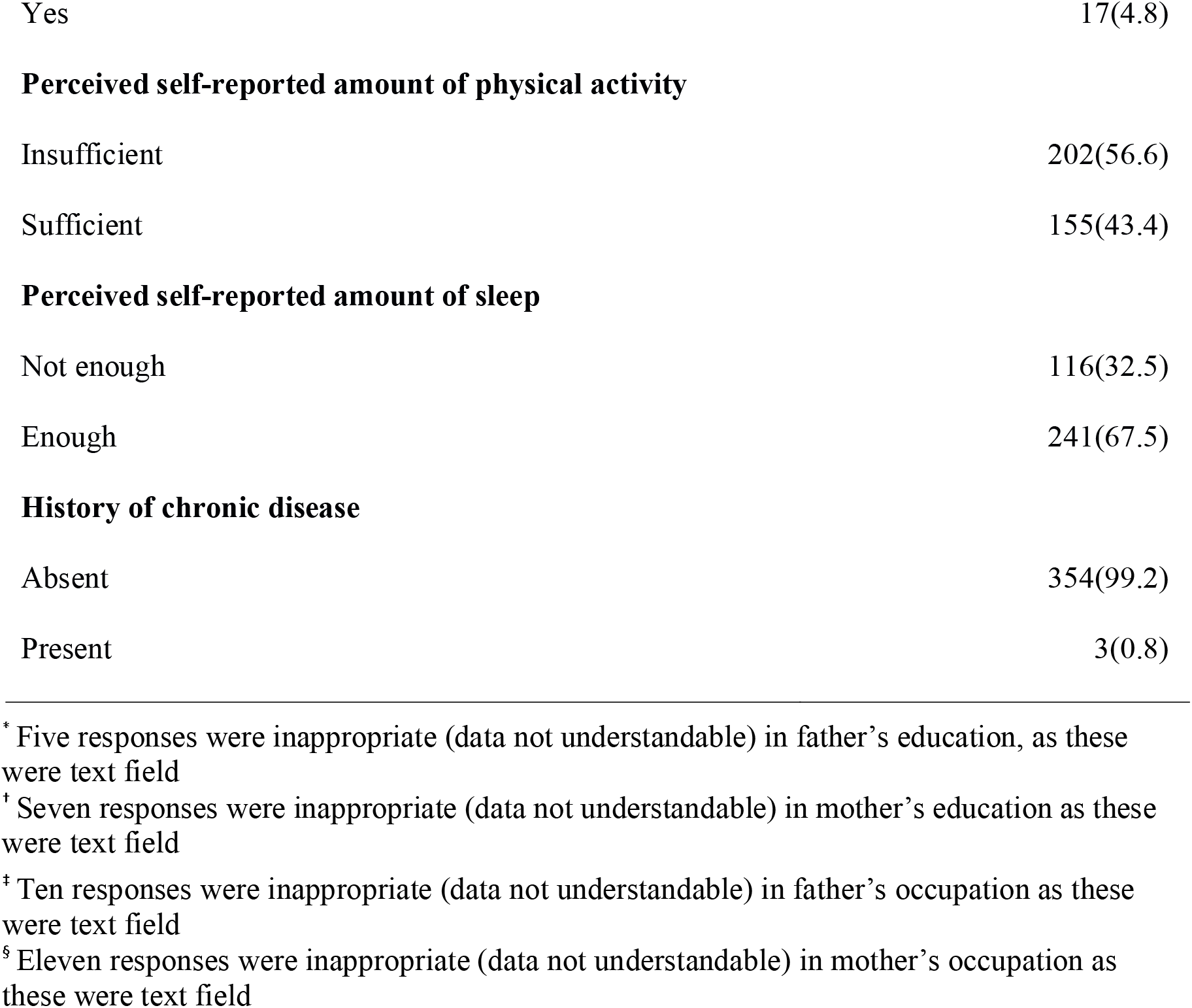
Socio-demographic and clinical characteristics of the study participants (n=357)

| Variable and category | N (%) |
| --- | --- |
| **Age (Years)** |  |
| 17-20 yrs | 267(74.8) |
| 21-25 yrs | 90(25.2) |
| **Gender** |  |
| Male | 256(71.7) |
| Female | 101(28.3) |
| **Permanent residence** |  |
| Urban | 260(72.8) |
| Rural | 97(27.2) |
| **Family type** |  |
| Nuclear | 306(85.7) |
| Extended | 51(14.3) |
| **Father's education (N=352)\*** |  |
| Less than High school | 36(10.1) |
| High School | 44(12.3) |
| Graduate | 147(41.2) |
| Post-graduate | 125(35) |
| **Mother's education (N=350)†** |  |
| Less than high school | 93(26.1) |

\* Five responses were inappropriate (data not understandable) in father's education, as these were text field
† Seven responses were inappropriate (data not understandable) in mother's education as these were text field

|  |  |
| --- | --- |
| High School | 45(12.6) |
| Graduation | 134(37.5) |
| Postgraduate | 78(21.8) |
| **Father's occupation (N=347)<sup>‡</sup>** |  |
| Unemployed | 10(2.8) |
| Semi-skilled, skilled, unskilled work | 16(4.5) |
| Clerical/shop | 112(31.4) |
| Semi-profession | 113(31.6) |
| Profession | 96(26.9) |
| **Mother's occupation(N=346)<sup>§</sup>** |  |
| Homemaker | 263(73.7) |
| Employed | 93(26.1) |
| **Per capita income (in USD)** |  |
| 0-68.72 | 64(17.9) |
| 68.74-343.67 | 181(50.7) |
| >343.68 | 112(31.4) |
| **Current tobacco use (last month)** |  |
| No | 342(95.8) |
| Yes | 15(4.2) |
| **Current alcohol use (last month)** |  |
| No | 340(95.2) |
| Yes | 17(4.8) |
| **Perceived self-reported amount of physical activity** |  |
| Insufficient | 202(56.6) |
| Sufficient | 155(43.4) |
| **Perceived self-reported amount of sleep** |  |
| Not enough | 116(32.5) |
| Enough | 241(67.5) |
| **History of chronic disease** |  |
| Absent | 354(99.2) |
| Present | 3(0.8) |

Out of 357, 111 (31.1%) participants had never heard the term ‘Sugar Sweetened Beverage’ previously. On asking what they understood by SSBs (question was asked before giving them the definition of SSBs), out of 357, their responses ranged from ‘don’t know’ in 26 (7.3%) cases, to a maximum of five SSBs. Participants either mentioned the type of drink or a name of the brand of SSB. Among them, 297 (83.2%) had understood soft drinks as SSBs, 80 (22.4%) as energy/sport drinks, 176 (47.3%) as packaged fruit drinks, 138 (38.7%) mentioned packaged milk product-based drinks (such as ‘*Lassi’*) and milkshakes, while only two (0.6%) participants mentioned other types of beverages (alcoholic beverages, honey water etc).

The lifetime prevalence of SSB consumption was 92.7% (331 participants). The current prevalence was 90.5 % as 323 had consumed SSBs in the last month. Median (IQR) consumption of SSBs was found to be 0.5 (0.2-1) L/participant/month, ranging from nil to 15 L/participant/month. Median (IQR) expenditure on SSBs in the last month was 1.37 (0.55-3.44) USD, ranging from nil to 137.47 USD. Average expenditure for buying SSBs in the last month was found to be 3.66% of participant’s per capita income. Most of the participants i.e. 253 out of 331 (76.4%) participants had consumed at least one of the SSBs within the last week (Figure 1). Also, out of those who had consumed SSBs, most participants i.e. 178 (53.8%) preferred soft drinks (Figure 2).

**Figure 1.**
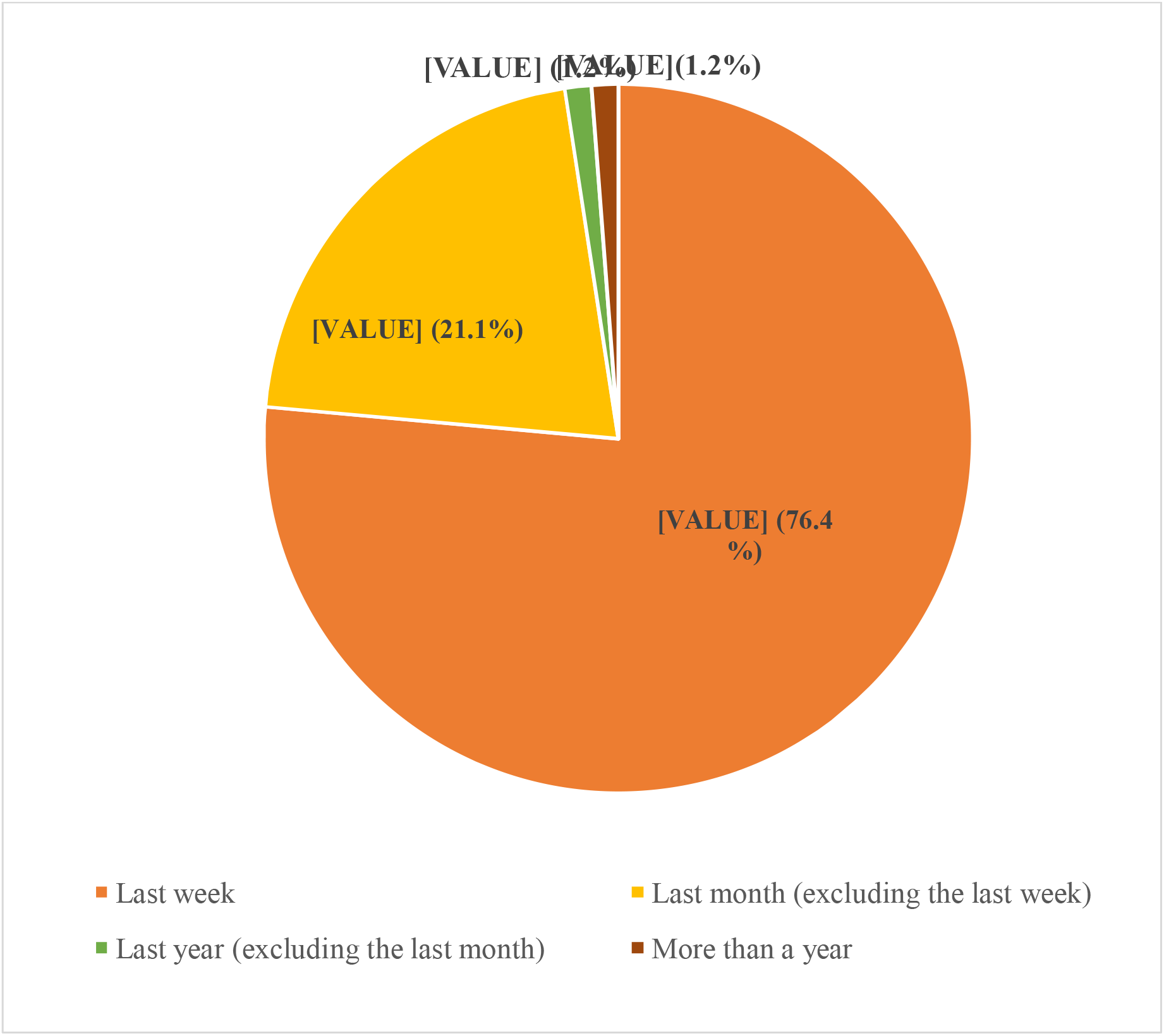
Frequency of SSB consumption among the study participants (N=331)

**Figure 2.**
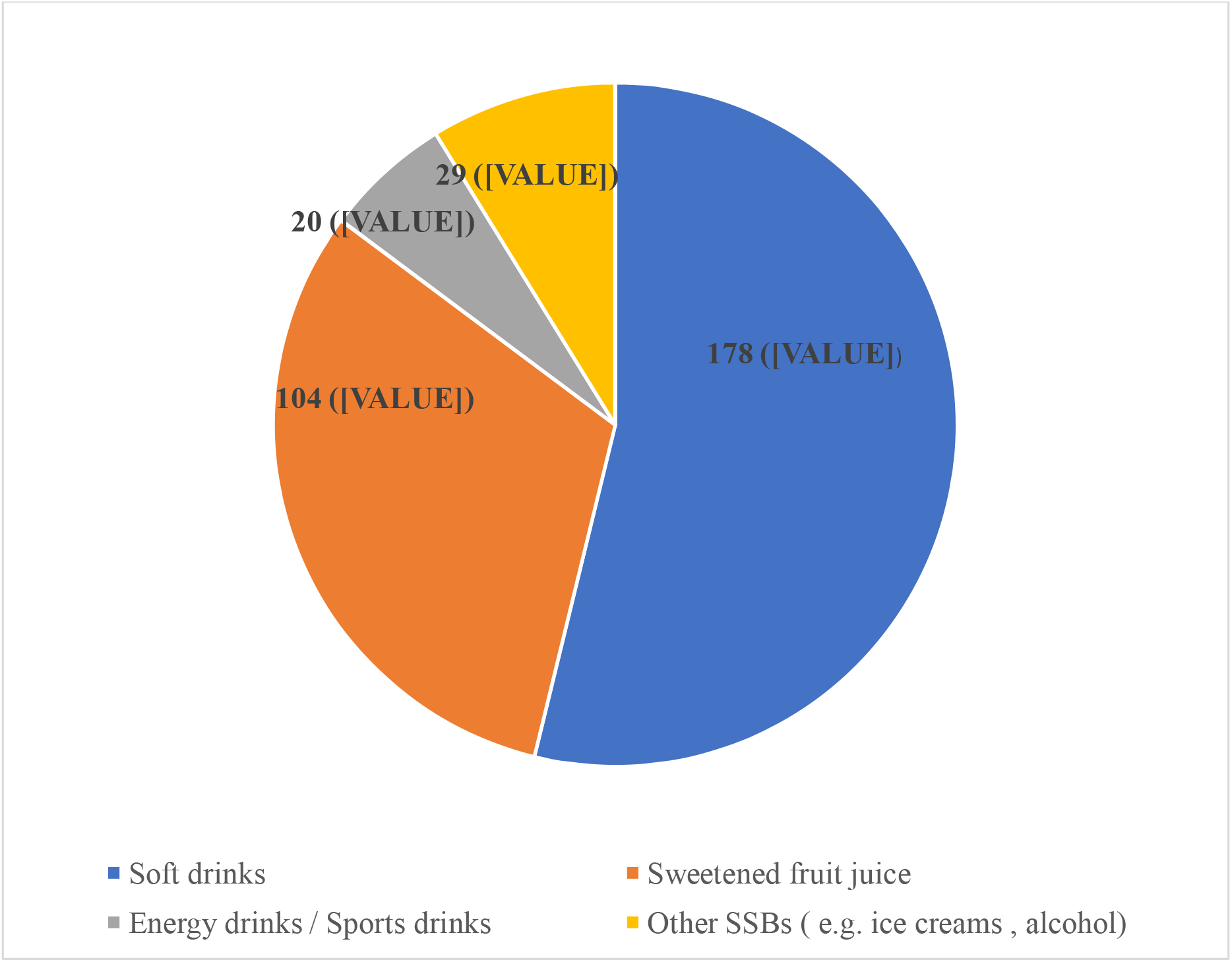
Preference of participants with respect to type of sugar sweetened beverage consumed (N=331)

Table 2 and 3 describe the results of univariable logistic regression analysis to find out the demographic, clinical and behavioural factors (related to consumption) predicting high intake of SSBs among the participants. Those having a permanent residence in an urban area had 1.7 times higher odds of high SSB intake as compared to those hailing from rural areas (*P = 0*.*034*). Also, those who had consumed SSB in the last week (more recent use) had 6.7 times higher odds of high SSB intake as compared to those who had not consumed SSB within the last one week (*P = 0*.*001*).

**Table 2:**
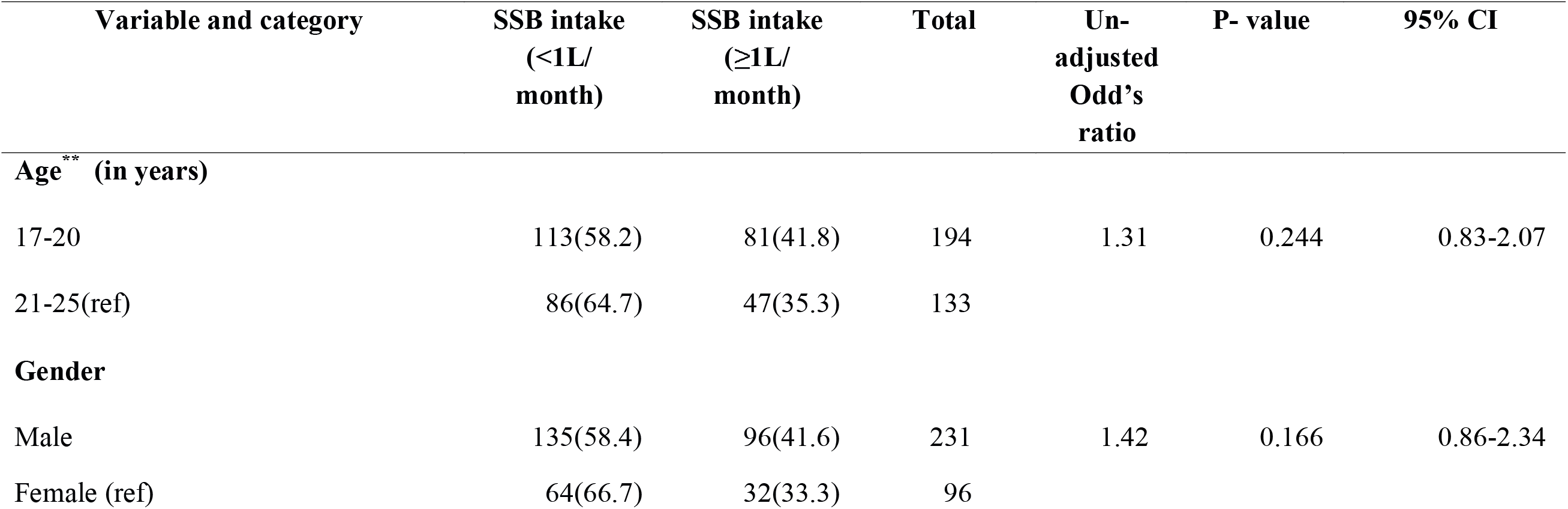
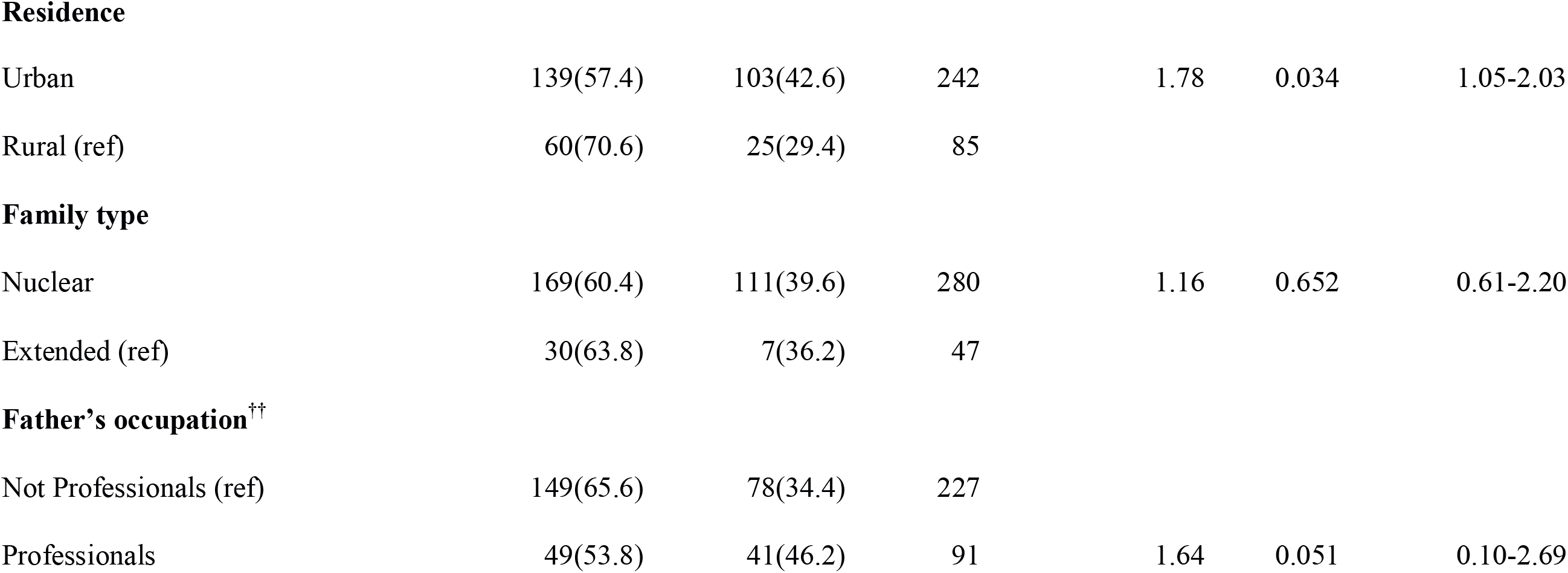
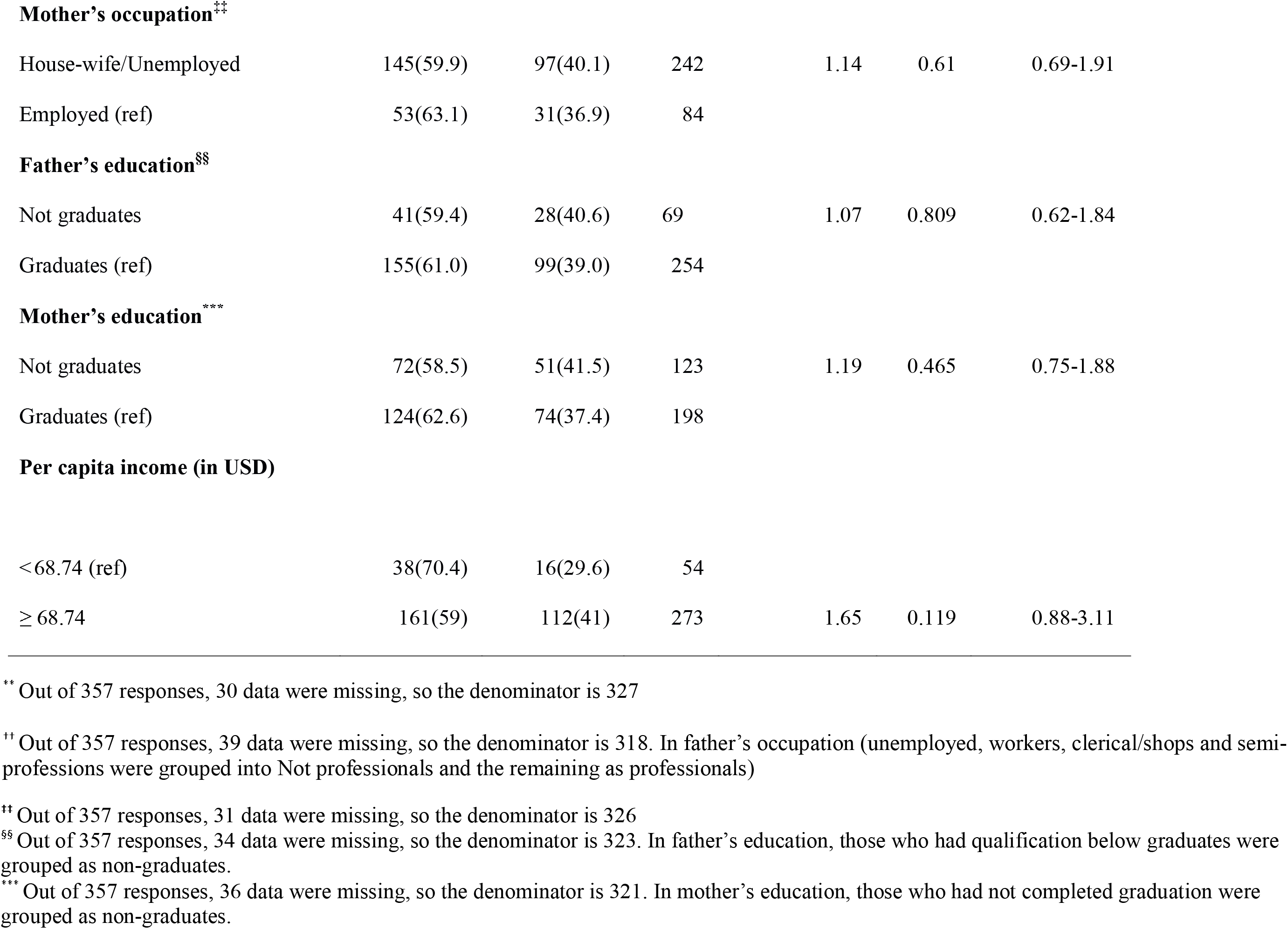
Univariable logistic regression analysis to determine the socio-demographic factors predicting participants’ high SSB intake (n=357)

| **Variable and category** | **SSB intake (<1L/month)** | **SSB intake (≥1L/month)** | **Total** | **Un-adjusted Odd's ratio** | **P- value** | **95% CI** |
| --- | --- | --- | --- | --- | --- | --- |
| **Age\*\* (in years)** |  |  |  |  |  |  |
| 17-20 | 113(58.2) | 81(41.8) | 194 | 1.31 | 0.244 | 0.83-2.07 |
| 21-25(ref) | 86(64.7) | 47(35.3) | 133 |  |  |  |
| **Gender** |  |  |  |  |  |  |
| Male | 135(58.4) | 96(41.6) | 231 | 1.42 | 0.166 | 0.86-2.34 |
| Female (ref) | 64(66.7) | 32(33.3) | 96 |  |  |  |

\*\* Out of 357 responses, 30 data were missing, so the denominator is 327

**Residence**
|  |  |  |  |  |  |  |
| --- | --- | --- | --- | --- | --- | --- |
| Urban | 139(57.4) | 103(42.6) | 242 | 1.78 | 0.034 | 1.05-2.03 |
| Rural (ref) | 60(70.6) | 25(29.4) | 85 |  |  |  |
| **Family type** |  |  |  |  |  |  |
| Nuclear | 169(60.4) | 111(39.6) | 280 | 1.16 | 0.652 | 0.61-2.20 |
| Extended (ref) | 30(63.8) | 7(36.2) | 47 |  |  |  |
| **Father's occupation<sup>††</sup>** |  |  |  |  |  |  |
| Not Professionals (ref) | 149(65.6) | 78(34.4) | 227 |  |  |  |
| Professionals | 49(53.8) | 41(46.2) | 91 | 1.64 | 0.051 | 0.10-2.69 |
| **Mother's occupation<sup>††</sup>** |  |  |  |  |  |  |
| House-wife/Unemployed | 145(59.9) | 97(40.1) | 242 | 1.14 | 0.61 | 0.69-1.91 |
| Employed (ref) | 53(63.1) | 31(36.9) | 84 |  |  |  |
| **Father's education<sup>§§</sup>** |  |  |  |  |  |  |
| Not graduates | 41(59.4) | 28(40.6) | 69 | 1.07 | 0.809 | 0.62-1.84 |
| Graduates (ref) | 155(61.0) | 99(39.0) | 254 |  |  |  |
| **Mother's education<sup>\*\*\*</sup>** |  |  |  |  |  |  |
| Not graduates | 72(58.5) | 51(41.5) | 123 | 1.19 | 0.465 | 0.75-1.88 |
| Graduates (ref) | 124(62.6) | 74(37.4) | 198 |  |  |  |
| **Per capita income (in USD)** |  |  |  |  |  |  |
| < 68.74 (ref) | 38(70.4) | 16(29.6) | 54 |  |  |  |
| ≥ 68.74 | 161(59) | 112(41) | 273 | 1.65 | 0.119 | 0.88-3.11 |

<sup>††</sup> Out of 357 responses, 31 data were missing, so the denominator is 326
<sup>§§</sup> Out of 357 responses, 34 data were missing, so the denominator is 323. In father's education, those who had qualification below graduates were grouped as non-graduates.
<sup>\*\*\*</sup> Out of 357 responses, 36 data were missing, so the denominator is 321. In mother's education, those who had not completed graduation were grouped as non-graduates.

**Table 3.**
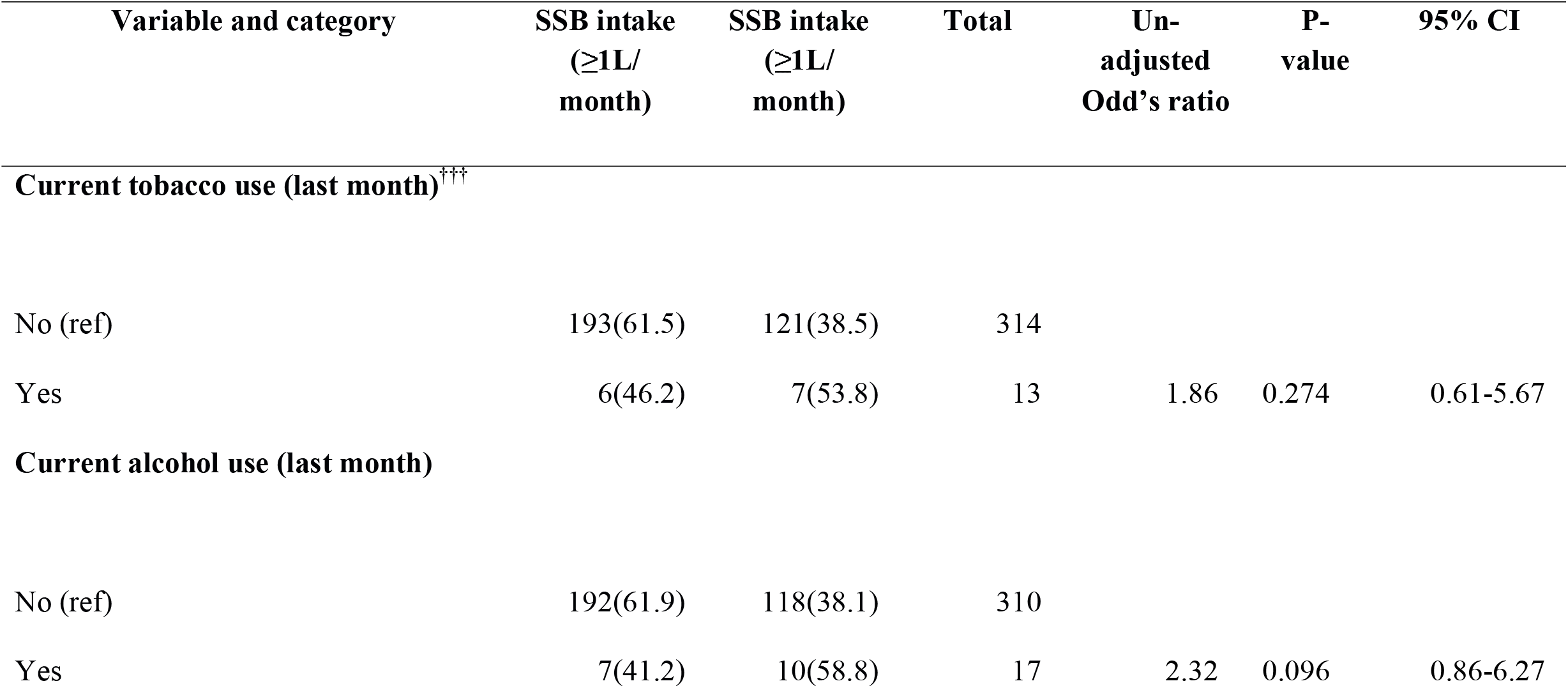
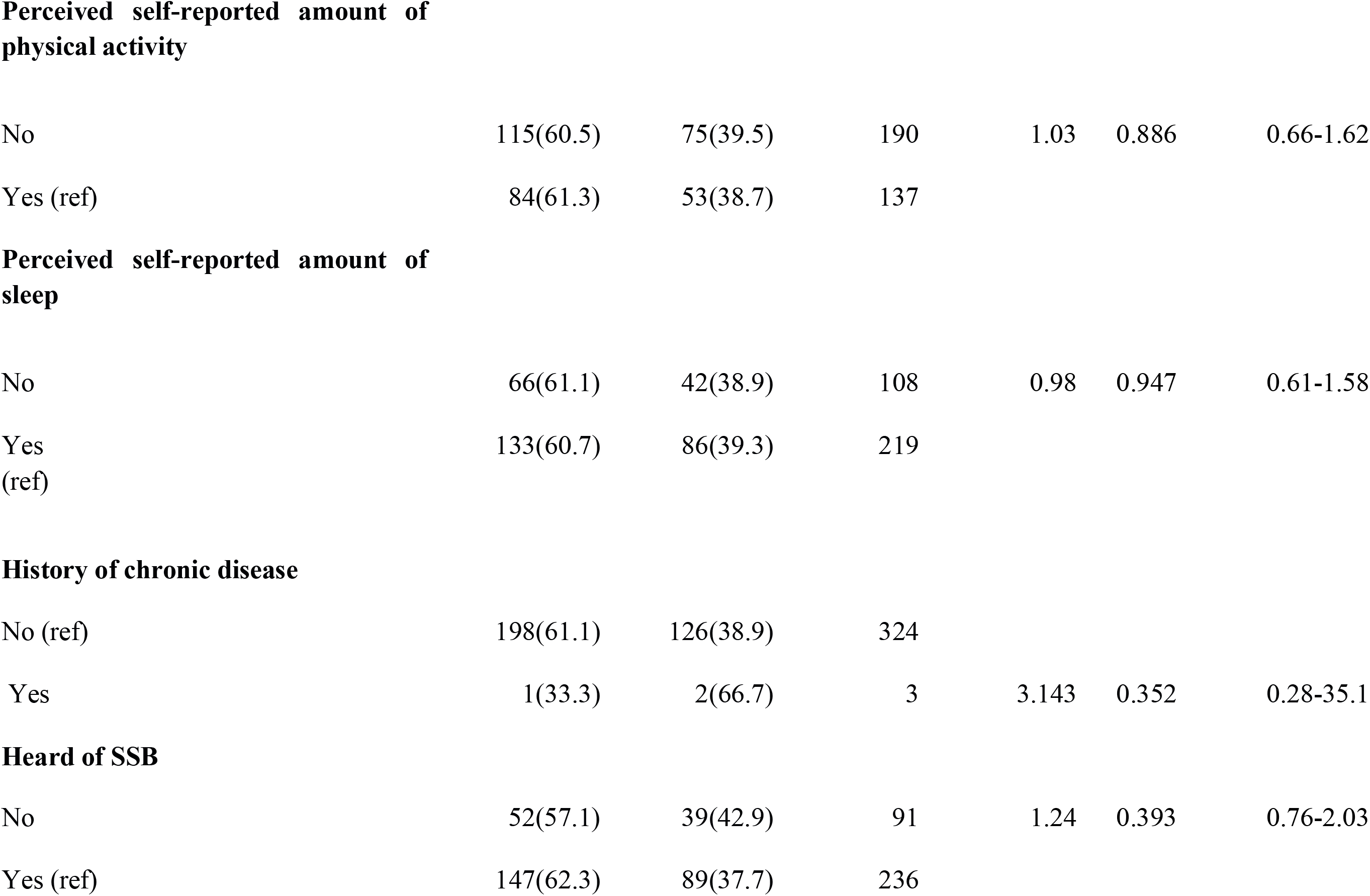
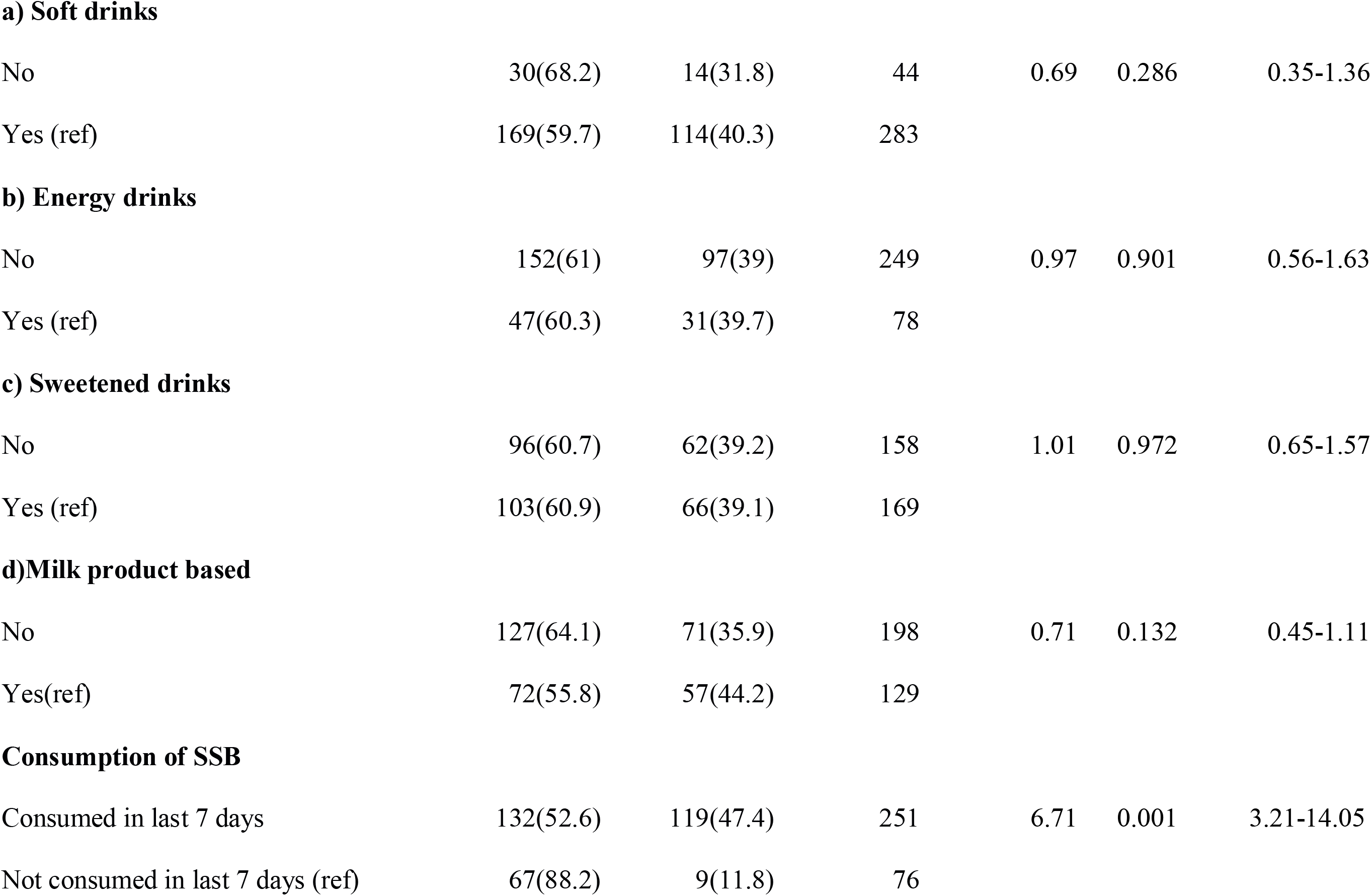
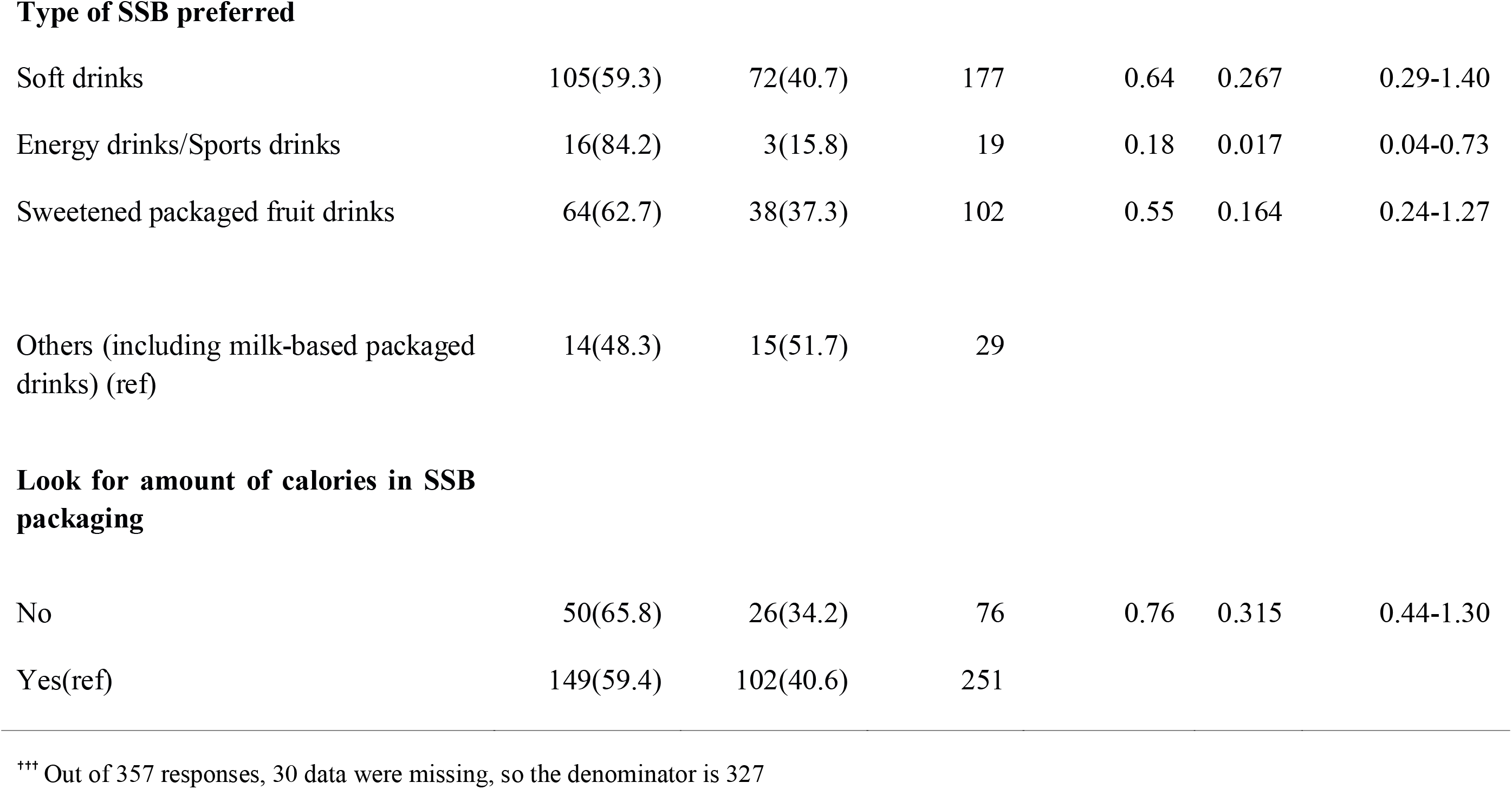
Univariable logistic regression analysis to determine the clinical and SSB consumption behavioural factors predicting participants’ high SSB intake (n=357)

| Variable and category | SSB intake<br>(≥1L/<br>month) | SSB intake<br>(≥1L/<br>month) | Total | Un-adjusted<br>Odd's ratio | P-value | 95% CI |
| --- | --- | --- | --- | --- | --- | --- |
| **Current tobacco use (last month)<sup>†††</sup>** |  |  |  |  |  |  |
| No (ref) | 193(61.5) | 121(38.5) | 314 |  |  |  |
| Yes | 6(46.2) | 7(53.8) | 13 | 1.86 | 0.274 | 0.61-5.67 |
| **Current alcohol use (last month)** |  |  |  |  |  |  |
| No (ref) | 192(61.9) | 118(38.1) | 310 |  |  |  |
| Yes | 7(41.2) | 10(58.8) | 17 | 2.32 | 0.096 | 0.86-6.27 |

<sup>†††</sup> Out of 357 responses, 30 data were missing, so the denominator is 327

**Perceived self-reported amount of physical activity**
|  |  |  |  |  |  |  |
| --- | --- | --- | --- | --- | --- | --- |
| No | 115(60.5) | 75(39.5) | 190 | 1.03 | 0.886 | 0.66-1.62 |
| Yes (ref) | 84(61.3) | 53(38.7) | 137 |  |  |  |
| **Perceived self-reported amount of sleep** |  |  |  |  |  |  |
| No | 66(61.1) | 42(38.9) | 108 | 0.98 | 0.947 | 0.61-1.58 |
| Yes (ref) | 133(60.7) | 86(39.3) | 219 |  |  |  |
| **History of chronic disease** |  |  |  |  |  |  |
| No (ref) | 198(61.1) | 126(38.9) | 324 |  |  |  |
| Yes | 1(33.3) | 2(66.7) | 3 | 3.143 | 0.352 | 0.28-35.1 |
| **Heard of SSB** |  |  |  |  |  |  |
| No | 52(57.1) | 39(42.9) | 91 | 1.24 | 0.393 | 0.76-2.03 |
| Yes (ref) | 147(62.3) | 89(37.7) | 236 |  |  |  |
| **a) Soft drinks** |  |  |  |  |  |  |
| No | 30(68.2) | 14(31.8) | 44 | 0.69 | 0.286 | 0.35-1.36 |
| Yes (ref) | 169(59.7) | 114(40.3) | 283 |  |  |  |
| **b) Energy drinks** |  |  |  |  |  |  |
| No | 152(61) | 97(39) | 249 | 0.97 | 0.901 | 0.56-1.63 |
| Yes (ref) | 47(60.3) | 31(39.7) | 78 |  |  |  |
| **c) Sweetened drinks** |  |  |  |  |  |  |
| No | 96(60.7) | 62(39.2) | 158 | 1.01 | 0.972 | 0.65-1.57 |
| Yes (ref) | 103(60.9) | 66(39.1) | 169 |  |  |  |
| **d)Milk product based** |  |  |  |  |  |  |
| No | 127(64.1) | 71(35.9) | 198 | 0.71 | 0.132 | 0.45-1.11 |
| Yes(ref) | 72(55.8) | 57(44.2) | 129 |  |  |  |
| **Consumption of SSB** |  |  |  |  |  |  |
| Consumed in last 7 days | 132(52.6) | 119(47.4) | 251 | 6.71 | 0.001 | 3.21-14.05 |
| Not consumed in last 7 days (ref) | 67(88.2) | 9(11.8) | 76 |  |  |  |
| **Type of SSB preferred** |  |  |  |  |  |  |
| Soft drinks | 105(59.3) | 72(40.7) | 177 | 0.64 | 0.267 | 0.29-1.40 |
| Energy drinks/Sports drinks | 16(84.2) | 3(15.8) | 19 | 0.18 | 0.017 | 0.04-0.73 |
| Sweetened packaged fruit drinks | 64(62.7) | 38(37.3) | 102 | 0.55 | 0.164 | 0.24-1.27 |
| Others (including milk-based packaged drinks) (ref) | 14(48.3) | 15(51.7) | 29 |  |  |  |
| **Look for amount of calories in SSB packaging** |  |  |  |  |  |  |
| No | 50(65.8) | 26(34.2) | 76 | 0.76 | 0.315 | 0.44-1.30 |
| Yes(ref) | 149(59.4) | 102(40.6) | 251 |  |  |  |

Table 4 describes the results of multivariable logistic regression analysis to determine the factors independently predicting high SSB intake among the participants. The variables whose P value were <0.25 in the univariable analysis were further considered for multivariable analysis, and these included age, gender, residence of the participants, father’s occupation, alcohol intake, per capita income, last consumption of SSB and type of SSB preferred. It was found that males had about 1.8 times higher odds of high SSB intake as compared to females (*P=0*.*04*). Those who had current alcohol consumption had about 4.1 times higher odds of high levels of SSB intake as compared to those who had not currently consumed alcohol *(P=0*.*02*). Similarly, those who had consumed a SSB in the last week had about 7.4 times higher odds of high SSB intake as compared to those who did not consume in the last week (*P=0*.*001*). However, those who preferred to consume energy drinks/sports drinks had low amount of SSB intake (adjusted Odds Ratio: 0.1, *P= 0*.*004*) as compared to those who had consumed other drinks such as milk product-based packaged drinks.

**Table 4:** Multivariable logistic regression analysis to determine factors independently predicting high intake (≥1L/month) of SSBs among the participants.

| Variable and category | Adjusted<br>Odd's ratio | P-value | 95% CI |
| --- | --- | --- | --- |
| **Gender** |  |  |  |
| Male | 1.83 | 0.040 | 1.03-3.25 |
| Female(ref) | - | - | - |
| **Current alcohol use** |  |  |  |
| No(ref) | - | - | - |
| Yes | 4.09 | 0.020 | 1.25-13.42 |
| **Last consumption of SSB** |  |  |  |
| Consumed in last 7 days | 7.36 | 0.001 | 3.41-15.87 |
| Not consumed in last 7 days(ref) | - | - | - |
| **Type of SSB preferred** |  |  |  |
| Soft drinks | 0.57 | 0.190 | 0.24-1.33 |
| Energy drinks/Sports drinks | 0.10 | 0.004 | 0.02-0.47 |
| Sweetened packaged fruit drinks | 0.55 | 0.193 | 0.23-1.35 |
| Others (including milk-based packaged drinks) (ref) | - | - | - |

## Discussion

The findings of our study are discussed below.

First, we found that the current (last one month) and lifetime prevalence of SSB consumption were 90.5% and 92.7% respectively. The reported prevalence of SSB consumption among Chinese children and adolescents aged 6-17 years was around 67%, which is lower than our study.(Gui *et al*., 2017) But a study conducted in Ontario showed that about 81.4% of the adolescents aged 11-20 years had consumed an SSB in the last week, and another 12% had consumed energy drinks.(Sampasa-Kanyinga, Hamilton and Chaput, 2018) This finding is similar to our study. The lower prevalence in Chinese adolescents can be ascribed to the lower age range, when parental supervision is usually high. We found that the median (IQR) consumption of SSBs was found to be 0.5 (0.2-1) L/ participant/month. Chinese adolescents had consumed around 0.61 servings per day (around 1.1 L/week).(Gui *et al*., 2017) Whereas, a study from USA reported much higher consumption of SSBs among adolescents i.e. 4.5L/week.(Wattelez *et al*., 2019) High intake of SSBs can be due to many factors such as lack of legislative measures, proper social message, peer pressure, cultural food habits etc. Also, intelligent and aggressive marketing strategies by SSB manufacturing companies play a part.(Skeie, Sandvær and Grimnes, 2019)

Second, we found that 49.9% of the participants preferred soft drinks, followed by 29.1% who preferred sweetened packaged fruit juice. Similarly, Park et al. found that students consumed more of soft drinks than energy drinks etc. Most (37%) of their participants preferred regular soda.(Park *et al*., 2012b) They attributed these findings to the frequent use of fast food restaurants and prolonged television viewing. About 50% of the participants in the study by Wang et al did consume carbonated SSBs.(Wang, Bleich and Gortmaker, 2008)

Third, we found that as compared to females, males had 1.8 times higher intake of SSBs. This fact has been reported in several studies.(Bjelland *et al*., 2011; Park *et al*., 2012a; Skeie, Sandvær and Grimnes, 2019) For instance, the adjusted odds ratios (OR) reported for males was 3.74 and 1.66 by Skeie G et al. and Park et al. respectively.(Park *et al*., 2012b; Skeie, Sandvær and Grimnes, 2019) These studies were not conducted on medical students though. One possible reason might be that male medical students in India are usually outdoors for longer times as compared to females, due to cultural and safety reasons, which increases their chances of buying packaged SSBs.

Fourth, we found that, the participants, who had consumed SSBs in last week, had 7.4 times high intake of SSBs. This may be attributed to negative symptoms experienced by participants by temporary stoppage/decrease in consumption, which might have lead to an urge to have higher and higher amounts of SSBs due to its addictive tendency. Falbe et al has reported that participants experienced headache, decreased satisfaction, low motivation to work and inability to concentrate during SSB cessation phase.(Falbe *et al*., 2019)

Fifth, participants who currently consumed alcohol had higher odds of high SSB intake than those not consuming alcohol. Some studies had contrasting findings, the reason ascribed being that nondrinkers might prefer SSBs to alcohol.(Park *et al*., 2014) Bleich et al too did find similar results.(Bleich *et al*., 2009) Cultural differences in type of alcohol preferences may partly explain our finding.

Sixth, participants who preferred energy drinks/sports drinks were less likely to consume high amounts of SSBs as compared to those consuming other type of SSBs. This may be because of the way they are marketed/advertised as elicited by a study of Bogart et al, with increase in advertisements resulting in increased consumption of SSBs.(Bogart *et al*., 2013)

Seventh, participants having permanent residence in an urban area consumed higher amount of SSBs than those from rural areas, although this was not found to be statistically significant in multivariable analysis. Some previous studies have reported similar findings.(Hearst, Pasch and Laska, 2012; Ramírez-Vélez, 2015; Mathur *et al*., 2020) However, one study from China did report that young children (aged 3-7 years) from rural area consumed more SSBs as compared to their urban counterparts.(Yu *et al*., 2016) The lower age group and higher economic status of Chinese may be a factor, it being an upper middle income country. Easy availability of different kinds of SSBs in urban areas and having a habitual urbanized lifestyle (unhealthy eating and drinking behaviors) as a part of their growing up, before getting into the medical course are possible factors.

Eighth, we did not find any statistically significant association between high intake of SSBs and age, family type, parent’s occupation and education, per capita income, tobacco use, physical activity, sleep and history of chronic disease. Skeie et al. too didn’t find any association between physical activity and SSBs consumed.(Skeie, Sandvær and Grimnes, 2019) Kenney et al. however, reported that increased SSB intake was associated with reduced physical activity.(Kenney and Gortmaker, 2017) One study reported poor sleep quality with higher intake of SSBs.(Park *et al*., 2012b)

So far, only few Indian studies have reported about SSB consumption. One study used secondary data from general population survey using only a limited number of variables.(Mathur *et al*., 2020) Also, that study did not estimate the actual quantity of SSB consumed. It has been reported earlier that medical students are prone to develop NCDs.(Adams-Campbell *et al*., 1988; Bertsias *et al*., 2003; Aslam, Mahmud and Waheed, 2004; Nyombi *et al*., 2016) To the best of our knowledge, this is the first study globally, reporting SSB consumption patterns and associated factors among medical students. Our sample size was also good. As data on prevalence of SSB use among medical students were not available, we did not do apriori sample size calculation. However, taking 50% prevalence (as in pilot studies), 20% relative precision and α = 0.05, the minimum sample size required was 100.

Since, the question on amount of SSB consumed in the last month was an open-ended question, there could be an element of reporting bias. It is possible, that some would have entered their consumption in litres, even after instructing them to fill in millilitres, leading to underreporting. Our findings might not be fully generalizable to private medical institutes, where the socio-economic status of students and campus culture may be different. We did not assess other unhealthy dietary practices, for example fast food intake, which might also influence SSB intake. Also, we could not objectively assess sedentary behaviour.

Targeted behavioural health interventions should be given to male medical students, alcohol users and frequent consumers of SSBs. Individual counselling by psychologists/psychiatrists should be arranged by the administration of the respective institutes for high SSB consumers. Students should be encouraged to consume home-made healthy drinks and abstain from taking alcohol. Legislation banning sale of SSBs within a certain radius of educational institutions should be brought in.

The prevalence of SSB consumption among medical students was high. Nearly half of the students preferred soft drinks, followed by sweetened packaged fruit juice (29%). Male gender, alcohol use in the last month and recent consumption of SSBs predicted high intake of SSBs. Behavioural change and legislative measures will help not only safeguard optimal health for future healthcare providers, but will also help prevent manpower shortage, excessive costs and compromised quality of care leading to inefficient health systems

## Data Availability

Will be attached as Supplementary File 1 Data sheet

## Supplementary Files

1. Supplementary File 1 – Data Sheet
2. Supplementary File 2 – STROBE checklist

## References

Adams-Campbell, L. L. et al. (1988) ‘Assessment of cardiovascular risk factors in Nigerian students.’, Arteriosclerosis: An Official Journal of the American Heart Association, Inc., 8(6), pp. 793–796. doi: 10.1161/01.ATV.8.6.793.

Aslam, F., Mahmud, H. and Waheed, A. (2004) ‘Cardiovascular health - Behaviour of medical students in Karachi’, JPMA. The Journal of the Pakistan Medical Association, 54, pp. 492–5.

Basu, S. et al. (2014) ‘Averting Obesity and Type 2 Diabetes in India through Sugar-Sweetened Beverage Taxation: An Economic-Epidemiologic Modeling Study’, PLoS Medicine. edited by T. Blakely, 11(1), p. e1001582. doi: 10.1371/journal.pmed.1001582.

Bertsias, G. et al. (2003) ‘Overweight and obesity in relation to cardiovascular disease risk factors among medical students in Crete, Greece’, BMC Public Health, 3, p. 3. doi: 10.1186/1471-2458-3-3.

Bjelland, M. et al. (2011) ‘Intakes and perceived home availability of sugar-sweetened beverages, fruit and vegetables as reported by mothers, fathers and adolescents in the HEIA (HEalth In Adolescents) study’, Public Health Nutrition, 14(12), pp. 2156–2165. doi: 10.1017/S1368980011000917.

Bleich, S. N. et al. (2009) ‘Increasing consumption of sugar-sweetened beverages among US adults: 1988-1994 to 1999-2004’, The American Journal of Clinical Nutrition, 89(1), pp. 372–381. doi: 10.3945/ajcn.2008.26883.

Bogart, L. M. et al. (2013) ‘Parental and Home Environmental Facilitators of Sugar-Sweetened Beverage Consumption Among Overweight and Obese Latino Youth’, Academic Pediatrics, 13(4), pp. 348–355. doi: 10.1016/j.acap.2013.02.009.

CDC (2018) Sugar Sweetened Beverage Intake, Centers for Disease Control and Prevention. Available at: https://www.cdc.gov/nutrition/data-statistics/sugar-sweetened-beverages-intake.html (Accessed: 17 March 2019).

Falbe, J. et al. (2019) ‘Potentially Addictive Properties of Sugar-Sweetened Beverages among Adolescents’, Appetite, 133, pp. 130–137. doi: 10.1016/j.appet.2018.10.032.

Gui, Z.-H. et al. (2017) ‘Sugar-Sweetened Beverage Consumption and Risks of Obesity and Hypertension in Chinese Children and Adolescents: A National Cross-Sectional Analysis’, Nutrients, 9(12). doi: 10.3390/nu9121302.

Hearst, M. O., Pasch, K. E. and Laska, M. N. (2012) ‘Urban versus suburban perceptions of the neighborhood food environment as correlates of adolescent food purchasing’, Public health nutrition, 15(2), pp. 299–306. doi: 10.1017/S1368980011002114.

Kenney, E. L. and Gortmaker, S. L. (2017) ‘United States Adolescents’ Television, Computer, Videogame, Smartphone, and Tablet Use: Associations with Sugary Drinks, Sleep, Physical Activity, and Obesity’, The Journal of Pediatrics, 182, pp. 144–149. doi: 10.1016/j.jpeds.2016.11.015.

KoBoToolbox | Data Collection Tools for Challenging Environments (no date) KoBoToolbox. Available at: https://kobotoolbox.org/ (Accessed: 17 July 2020).

Malik, V. S. et al. (2010) ‘Sugar-Sweetened Beverages and Risk of Metabolic Syndrome and Type 2 Diabetes’, Diabetes Care, 33(11), pp. 2477–2483. doi: 10.2337/dc10-1079.

Malik, V. S., Schulze, M. B. and Hu, F. B. (2006) ‘Intake of sugar-sweetened beverages and weight gain: a systematic review’, The American Journal of Clinical Nutrition, 84(2), pp. 274–288. doi: 10.1093/ajcn/84.1.274.

Mathur, M. R. et al. (2020) ‘Determinants of Sugar-Sweetened Beverage Consumption among Indian Adults: Findings from the National Family Health Survey-4’, Indian Journal of Community MedicineJ: Official Publication of Indian Association of Preventive & Social Medicine, 45(1), pp. 60–65. doi: 10.4103/ijcm.IJCM_201_19.

Nyombi, K. V. et al. (2016) ‘High prevalence of hypertension and cardiovascular disease risk factors among medical students at Makerere University College of Health Sciences, Kampala, Uganda’, BMC research notes, 9, p. 110. doi: 10.1186/s13104-016-1924-7.

Obesity and overweight (no date a). Available at: https://www.who.int/news-room/fact-sheets/detail/obesity-and-overweight (Accessed: 24 July 2021).

Obesity and overweight (no date b). Available at: https://www.who.int/news-room/fact-sheets/detail/obesity-and-overweight (Accessed: 30 May 2020).

Park, S. et al. (2012a) ‘Factors Associated with Sugar-Sweetened Beverage Intake among United States High School Students’, The Journal of nutrition, 142(2), pp. 306–312. doi: 10.3945/jn.111.148536.

Park, S. et al. (2012b) ‘Factors Associated with Sugar-Sweetened Beverage Intake among United States High School Students’, The Journal of nutrition, 142(2), pp. 306–312. doi: 10.3945/jn.111.148536.

Park, S. et al. (2014) ‘Consumption of Sugar-Sweetened Beverages Among US Adults in 6 States: Behavioral Risk Factor Surveillance System, 2011’, Preventing Chronic Disease, 11. doi: 10.5888/pcd11.130304.

Ramírez-Vélez, R. (2015) ‘DIFERENCIAS DEMOGRÁFICAS Y SOCIOECONÓMICAS ASOCIADAS AL CONSUMO DE’, NUTRICION HOSPITALARIA, (6), pp. 2479–2486. doi: 10.3305/nh.2015.31.6.8986.

Sampasa-Kanyinga, H., Hamilton, H. A. and Chaput, J.-P. (2018) ‘Sleep duration and consumption of sugar-sweetened beverages and energy drinks among adolescents’, Nutrition, 48, pp. 77–81. doi: 10.1016/j.nut.2017.11.013.

Singh, G. M. et al. (2015) ‘Global, Regional, and National Consumption of Sugar-Sweetened Beverages, Fruit Juices, and Milk: A Systematic Assessment of Beverage Intake in 187 Countries’, PLOS ONE. Edited by M. Müller, 10(8), p. e0124845. doi: 10.1371/journal.pone.0124845.

Skeie, G., Sandvær, V. and Grimnes, G. (2019) ‘Intake of Sugar-Sweetened Beverages in Adolescents from Troms, Norway—The Tromsø Study: Fit Futures’, Nutrients, 11(2), p. 211. doi: 10.3390/nu11020211.

Sohn, W., Burt, B. A. and Sowers, M. R. (2006) ‘Carbonated soft drinks and dental caries in the primary dentition’, Journal of Dental Research, 85(3), pp. 262–266. doi: 10.1177/154405910608500311.

Wang, Y. C., Bleich, S. N. and Gortmaker, S. L. (2008) ‘Increasing Caloric Contribution From Sugar-Sweetened Beverages and 100% Fruit Juices Among US Children and Adolescents, 1988-2004’, PEDIATRICS, 121(6), pp. e1604–e1614. doi: 10.1542/peds.2007-2834.

Wani, R. T. (2019) ‘Socioeconomic status scales-modified Kuppuswamy and Udai Pareekh’s scale updated for 2019’, Journal of Family Medicine and Primary Care, 8(6), p. 1846. doi: 10.4103/jfmpc.jfmpc_288_19.

Wattelez, G. et al. (2019) ‘Sugar-Sweetened Beverage Consumption and Associated Factors in School-Going Adolescents of New Caledonia’, Nutrients, 11(2). doi: 10.3390/nu11020452.

WHO | Reducing consumption of sugar-sweetened beverages to reduce the risk of childhood overweight and obesity (no date). Available at: https://www.who.int/elena/titles/ssbs_childhood_obesity/en/ (Accessed: 17 March 2019).

Yang, Q. et al. (2014) ‘Added sugar intake and cardiovascular diseases mortality among US adults’, JAMA internal medicine, 174(4), pp. 516–524. doi: 10.1001/jamainternmed.2013.13563.

Yu, P. et al. (2016) ‘Consumption of sugar-sweetened beverages and its association with overweight among young children from China’, Public Health Nutrition, 19(13), pp. 2336–2346. doi: 10.1017/S1368980016001373.

